# Bamlanivimab reduces nasopharyngeal SARS-CoV-2 RNA levels but not symptom duration in non-hospitalized adults with COVID-19

**DOI:** 10.1101/2021.12.17.21268009

**Authors:** Kara W. Chew, Carlee Moser, Eric S. Daar, David A. Wohl, Jonathan Z. Li, Robert Coombs, Justin Ritz, Mark Giganti, Arzhang Cyrus Javan, Yijia Li, Carlos Malvestutto, Paul Klekotka, Karen Price, Ajay Nirula, William Fischer, Veenu Bala, Ruy M. Ribeiro, Alan S. Perelson, Courtney V. Fletcher, Joseph J. Eron, Judith S. Currier, Michael D. Hughes, Davey M. Smith

## Abstract

**Importance:** The antiviral activity and efficacy of anti-SARS-CoV-2 monoclonal antibody (mAb) therapies to accelerate recovery from COVID-19 is important to define.

**Objective:** To determine safety and efficacy of the mAb bamlanivimab to reduce nasopharyngeal (NP) SARS-CoV-2 RNA levels and symptom duration.

**Design:** ACTIV-2/A5401 is a randomized, blinded, placebo-controlled platform trial. Two dose cohorts were enrolled between August 19 and November 17, 2020 for phase 2 evaluation: in the first, participants were randomized 1:1 to bamlanivimab 7000 mg versus placebo, and in the second to bamlanivimab 700 mg versus placebo. Randomization was stratified by time from symptom onset (≤ or >5 days) and risk of progression to severe COVID-19 (“higher” vs “lower”).

**Setting:** Multicenter trial conducted at U.S. sites.

**Participants:** Non-hospitalized adults ≥18 years of age with positive SARS-CoV-2 antigen or nucleic acid test within 7 days, ≤10 days of COVID-19 symptoms, and with oxygen saturation ≥92% within 48 hours prior to study entry.

**Intervention:** Single infusion of bamlanivimab (7000 or 700 mg) or placebo.

**Main Outcomes and Measures:** Detection of NP SARS-CoV-2 RNA at days 3, 7, 14, 21, and 28, time to improvement of all of 13 targeted COVID-19 symptoms by daily self-assessment through day 28, and grade 3 or higher treatment emergent adverse events (TEAEs) through day 28. Secondary measures included quantitative NP SARS-CoV-2 RNA, all-cause hospitalizations and deaths (composite), area under the curve of symptom scores from day 0 through day 28, plasma bamlanivimab concentrations, plasma and serum inflammatory biomarkers, and safety through week 24.

**Results:** Ninety-four participants were enrolled to the 7000 mg cohort and 223 to the 700 mg cohort and initiated study intervention. The proportion meeting protocol criteria for “higher” risk for COVID-19 progression was 42% and 51% for the 7000 and 700 mg cohort, respectively. Median time from symptom onset at study entry for both cohorts was 6 days. There was no difference in the proportion with undetectable NP SARS-CoV-2 RNA at any post-treatment timepoints (risk ratio compared to placebo, 0.82-1.05 for 7000 mg dose [overall p=0.88] and 0.81-1.21 for 700 mg dose [overall p=0.49]), time to symptom improvement (median of 21 vs 18.5 days, p=0.97, for 7000 mg bamlanivimab vs placebo and 24 vs 20.5 days, p=0.08, for 700 mg bamlanivimab vs placebo), or grade 3+ TEAEs with either dose compared to placebo. Median NP SARS-CoV-2 RNA levels were lower at day 3 and C-reactive protein, ferritin, and fibrinogen levels significantly reduced at days 7 and 14 for bamlanivimab 700 mg compared to placebo, with similar trends observed for bamlanivimab 7000 mg. Viral decay modeling supported more rapid decay with bamlanivimab compared to placebo.

**Conclusions and Relevance:** Treatment with bamlanivimab 7000 mg and 700 mg was safe and compared to placebo led to more rapid reductions in NP SARS-CoV-2 RNA and inflammatory biomarkers, but did not decrease time to symptom improvement. The clinical utility of mAbs for outcomes other than hospitalizations and deaths is uncertain.

**Trial Registration:** ClinicalTrials.gov Identifier: NCT04518410

**KEY POINTS:** *Question:* What is the safety and efficacy of bamlanivimab monoclonal antibody (mAb) treatment for mild to moderate COVID-19?

*Findings:* In this randomized, placebo-controlled phase 2 trial of 317 non-hospitalized adults with COVID-19, there was no relationship between symptoms or disease progression risk and nasopharyngeal (NP) virus shedding. Bamlanivimab was safe and reduced NP SARS-CoV-2 RNA levels and inflammatory biomarker levels more than placebo, but did not shorten symptom duration.

*Meaning:* Nasal virus shedding was not associated with symptoms or baseline risk factors for severe COVID-19. Bamlanivimab, which has been associated with reduced hospitalizations in high-risk individuals, demonstrated antiviral activity with early post-treatment NP sampling but did not accelerate symptom improvement. The clinical utility of bamlanivimab for outcomes other than hospitalizations and deaths, including longer-term outcomes, is uncertain.

## INTRODUCTION

Severe acute respiratory syndrome coronavirus 2 (SARS-CoV-2), the virus that causes Coronavirus disease 2019 (COVID-19), continues to exert an enormous global public health and economic toll, and in the U.S. case-fatality rates exceed estimates for the 1918 influenza pandemic.^1^ Anti-SARS-CoV-2 monoclonal antibody (mAb)-based therapies have shown sufficient clinical efficacy to receive emergency authorization (EUA) by regulatory agencies for the treatment of early COVID-19 in non-hospitalized persons.^2-5^

Bamlanivimab is a neutralizing immunoglobulin G (IgG)-1 mAb directed to the receptor binding domain (RBD) of the spike (S) protein of SARS-CoV-2. On November 9, 2020, based on data from the Blocking Viral Attachment and Cell Entry with SARS-CoV-2 Neutralizing Antibodies (BLAZE-1) trial (NCT04427501)^2^, the FDA issued an EUA for its use as a one-time 700 mg intravenous (IV) infusion for the treatment for mild to moderate COVID-19 in non-hospitalized adults with risk factors for progression to severe disease who were within 10 days of symptom onset. Since then, the emergence of SARS-CoV-2 variants with decreased susceptibility *in vitro* to bamlanivimab^6,7^ led to withdrawal of the EUA, although bamlanivimab in combination with etesevimab continues to have an active EUA for the treatment of non-hospitalized adults.

With the rapid development of additional anti-SARS-CoV-2 mAbs, understanding the antiviral activity and characterizing the clinical benefits of these agents remains a critical need. Here, we describe the safety, virologic, and clinical outcomes of a placebo-controlled phase 2 evaluation of bamlanivimab at two doses, 7000 mg and 700 mg, in non-hospitalized adults with COVID-19.

## METHODS

### Trial Design

The ACTIV-2/A5401 study is an ongoing multicenter phase 2/3 adaptive platform randomized controlled trial for the evaluation of therapeutics for early COVID-19 in non-hospitalized adults (see **Supplement 1** for the ACTIV-2/A5401 protocol). Phase 2 results for bamlanivimab compared to placebo are reported here. All participants for this phase 2 analysis were enrolled in the U.S., across 38 sites (listed in **Supplement 2**). The protocol was approved by a central institutional review board (IRB), Advarra (Pro00045266), with additional local IRB review and approval as required by participating sites. All participants provided written informed consent.

### Participants

Adults 18 years of age or older with documented SARS-CoV-2 infection by an FDA-authorized antigen or nucleic acid test from a sample collected within 7 days prior to anticipated study entry, no more than 10 days of COVID-19 symptoms at time of anticipated study entry, ongoing symptoms (not including loss of taste or smell) within 48 hours prior to study entry, resting peripheral oxygen saturation levels ≥92%, and without the need for hospitalization were eligible. Complete eligibility criteria are provided in the protocol in **Supplement 1**.

### Randomization

Participants were randomly assigned by a web-based interactive response system in a 1:1 ratio to receive either bamlanivimab or placebo. Randomization was stratified by time from symptom onset (≤ or >5 days) and risk of progression to severe COVID-19 (“higher” vs “lower”). “Higher” risk was defined in the protocol as meeting any of the following: age ≥55 years or having a comorbidity (chronic lung disease or moderate to severe asthma, body mass index >35 kg/m^2^, hypertension, cardiovascular disease, diabetes, or chronic kidney or liver disease).

### Study Intervention

An initial dose of 7000 mg of bamlanivimab was chosen for study based on pharmacokinetic (PK) and preliminary safety data. After phase 2 data on bamlanivimab 700 mg, 2800 mg, and 7000 mg from an outside trial did not conclusively demonstrate a dose-response effect of higher doses on declines in nasopharyngeal (NP) SARS-CoV-2 RNA levels, the protocol was amended to evaluate the 700 mg dose.^8^ Bamlanivimab or placebo (normal saline) was administered as a single IV infusion over approximately 60 minutes.

### Primary and Secondary Outcomes

The phase 2 study was designed to evaluate the safety of bamlanivimab and determine the efficacy of bamlanivimab to reduce the duration of COVID-19 symptoms and SARS-CoV-2 RNA shedding from NP swabs. NP swabs were collected on days 0 (day of study intervention, pre-intervention), 3, 7, 14, 21, and 28. Participants completed a study diary each day from day 0 to day 28, which included self-assessment of 13 targeted COVID-19 symptoms, scored by the participant as absent, mild, moderate, or severe (see **Supplement 2** for symptom diary). A numerical total symptom score was calculated for each day by summing scores for each symptom, with absent scored as 0, mild as 1, moderate as 2, and severe as 3; therefore, the range of total symptoms scores was 0 to 39.

Primary clinical outcome measures were: 1) development of a Grade 3 or higher treatment-emergent adverse event (TEAE) through 28 days; 2) detection (detectable versus undetectable) of SARS-CoV-2 RNA from NP swabs at days 3, 7, 14, 21, and 28; 3) duration of targeted COVID-19-associated symptoms from day 0 (utilizing daily diary data), where duration was defined as the number of days from day 0 to the last day on or before study day 28 when any targeted symptoms that were self-assessed as moderate or severe at day 0 (before study intervention) were still scored as moderate or severe (i.e., not mild or absent), or any targeted symptoms scored as mild or absent at day 0 were still scored as mild or worse (i.e. not absent). Participants with ongoing unimproved symptoms at day 28 were treated as having a symptom duration of 28 days for analysis.

Secondary outcome measures included all-cause hospitalization and death, quantitative NP SARS-CoV-2 RNA levels; area under the curve (AUC) of symptom scores from days 0-28; and change in inflammatory markers from baseline through week 24; development of AEs of special interest (AESIs, specifically Grade 1 or higher infusion-related reactions [IRRs] and Grade 1 or higher allergic/hypersensitivity reactions), and serious adverse events (SAEs) through day 28 and through week 24; and progression of 1 or more COVID-19-associated symptoms to a worse status than recorded in the study diary at entry. The full set of secondary and exploratory outcome measures are provided in **Supplement 1** (protocol version 1.0).

### Virology

NP and anterior nasal (AN) samples were collected using standardized swabs and collection procedures. AN swabs were self-collected by participants daily days 0-14. Site-collected NP swabs, AN swabs collected on site, and EDTA plasma samples were frozen and stored at -80°C (−65°C to -95°C) on the day of collection. AN swabs collected remotely were stored at cool temperatures (refrigerated or in a study-provided cooler with a combination of refrigerated and frozen gel packs) and returned to the site and frozen at -80°C (−65°C to -95°C) within 7 days of collection. Samples were shipped on dry ice to a central laboratory (University of Washington) for quantitative SARS-CoV-2 RNA testing using the Abbott m2000sp/rt platform with a validated internal standard.^9^ The collection, storage, processing, and assay methods have previously been validated.^9^ The assay limit of detection (LoD) was 1.4 log_10_ copies/mL, lower limit of quantification (LLoQ) was 2 log_10_ copies/mL, and upper limit of quantification (ULoQ) was 7 log_10_ copies/mL. For samples with RNA levels >ULoQ, the assay was rerun with dilutions to obtain a quantitative value.

The rates of decline of NP and AN virus after study entry were quantified in separate models using Monolix 2020 (Lixoft, Antony, France). Methods for model fitting and selection are described in **Supplement 2**.

### Serum and plasma biomarkers

Inflammatory and coagulation markers including lactate dehydrogenase (LDH), C-reactive protein (CRP), ferritin, D-dimer, prothrombin time (PT) and international normalized ratio (INR), partial thromboplastin time (PTT), and fibrinogen were measured in real time by a central clinical laboratory at days 0, 7, 14, 21, and 28 and weeks 12 and 24.

### Pharmacokinetic (PK) analysis

Blood samples for quantitation of bamlanivimab serum concentrations were collected pre-dose and at the following times after the end of infusion: 30 minutes, days 14 and 28 and weeks 12 and 24. PK parameters of interest were maximum concentration (Cmax), area under the concentration-time curve from time 0 to infinity (AUC_0-∞_), elimination half-life and total body clearance (CL), which were calculated based on the statistical moment theory using the trapezoidal rule and linear regression (WinNonLin, Certara, Princeton, NJ, USA).

### Power analysis and sample size calculation

The sample size of 110 participants randomized to each arm was selected to give high power to identify an active agent based on the primary virologic outcome. At the time the study was designed, there were no data in outpatients with COVID-19 to inform expected differences in proportion undetectable for NP SARS-CoV-2 RNA over 28 days. We estimated that a 20% absolute increase in the proportion undetectable would be clinically relevant, and 110 participants assigned to each arm would have 82.5 to 95.5% power dependent on the proportion undetectable in the placebo arm, with a two-sided Type I error rate of 5%.

### Statistical Analysis

The analysis population included all participants who initiated study intervention (bamlanivimab or placebo). Four participants enrolled to the 7000 mg dose cohort received 700 mg bamlanivimab or placebo and were included in the 700 mg analysis population (the randomization to active agent or placebo remained valid). One participant enrolled in the 700 mg dose cohort received 7000 mg bamlanivimab and was included in the 7000 mg analysis population.

The proportion of participants experiencing a grade 2 or higher and grade 3 or higher TEAE was compared between arms using log-binomial regression and summarized with a risk ratio (RR), corresponding 95% CI and p-value based on the Wald test. The proportion of participants with undetectable SARS-CoV-2 RNA was compared between arms across study visits using Poisson regression adjusted for baseline (day 0) log_10_ transformed SARS-CoV-2 RNA level and summarized with RR and 95% CI at each time, and Wald test across the multiple times. Quantitative SARS-CoV-2 RNA levels were compared between arms using Wilcoxon rank-sum tests, separately at each post-entry study visit, without adjustment for baseline value. For this comparison, results below the LoD were imputed as the lowest rank and values above the LoD but below the LLoQ were imputed as the second lowest rank. For summaries of quantitative RNA levels, values below the LoD were imputed as 0.7 log_10_ copies/ml (i.e., half the distance from zero to the LoD), values above the LoD but below the LLoQ were imputed as 1.7 log_10_ copies/ml (i.e., half the distance from the LoD to the LLoQ), and values above the ULoQ were imputed as 8 log_10_ copies/ml if a numerical value was not available. The two dose cohorts (700 and 7000 mg) were combined for analyses of baseline NP, AN, and plasma SARS-CoV-2 RNA levels. Spearman correlations evaluated associations between total symptoms scores and NP and AN RNA levels, Wilcoxon tests and chi-square tests were used to evaluate NP and AN RNA levels and symptom scores between subgroups.

Participant-specific symptom durations and area under the curve (AUC) of total symptom score from days 0-28 were compared between arms using a Wilcoxon rank sum test. Due to the small number of hospitalization/death events, the proportion hospitalized/dead in the bamlanivimab and placebo arms was summarized with descriptive statistics and compared between arms using Fisher’s exact test as a post-hoc analysis. Change from baseline in log-transformed inflammatory and coagulation biomarker levels were compared between bamanivimab and placebo arms using Wilcoxon tests.

No adjustment was made for the multiple comparisons across outcome measures. Statistical analyses were conducted using SAS version 9.4 and R version 4.1.0. See **Supplement 1** for complete Statistical Analysis Plan.

## RESULTS

### Characteristics of Participants and Retention in Follow-up

The analysis population included 94 participants in the bamlanivimab 7000 mg dose cohort (48 bamlanivimab, 46 placebo) enrolled between August 19 and November 15, 2020, and 223 participants in the bamlanivimab 700 mg dose cohort (111 bamlanivimab, 112 placebo) enrolled between October 12 and November 17, 2020. Across bamlanivimab and placebo arms, 3 (3.2%) of the 7000 mg group, and 6 (2.6%) of the 700 mg group prematurely discontinued the study prior to day 28 (**Figure 1**, Consort Flow Diagrams).

**Figure 1A.**
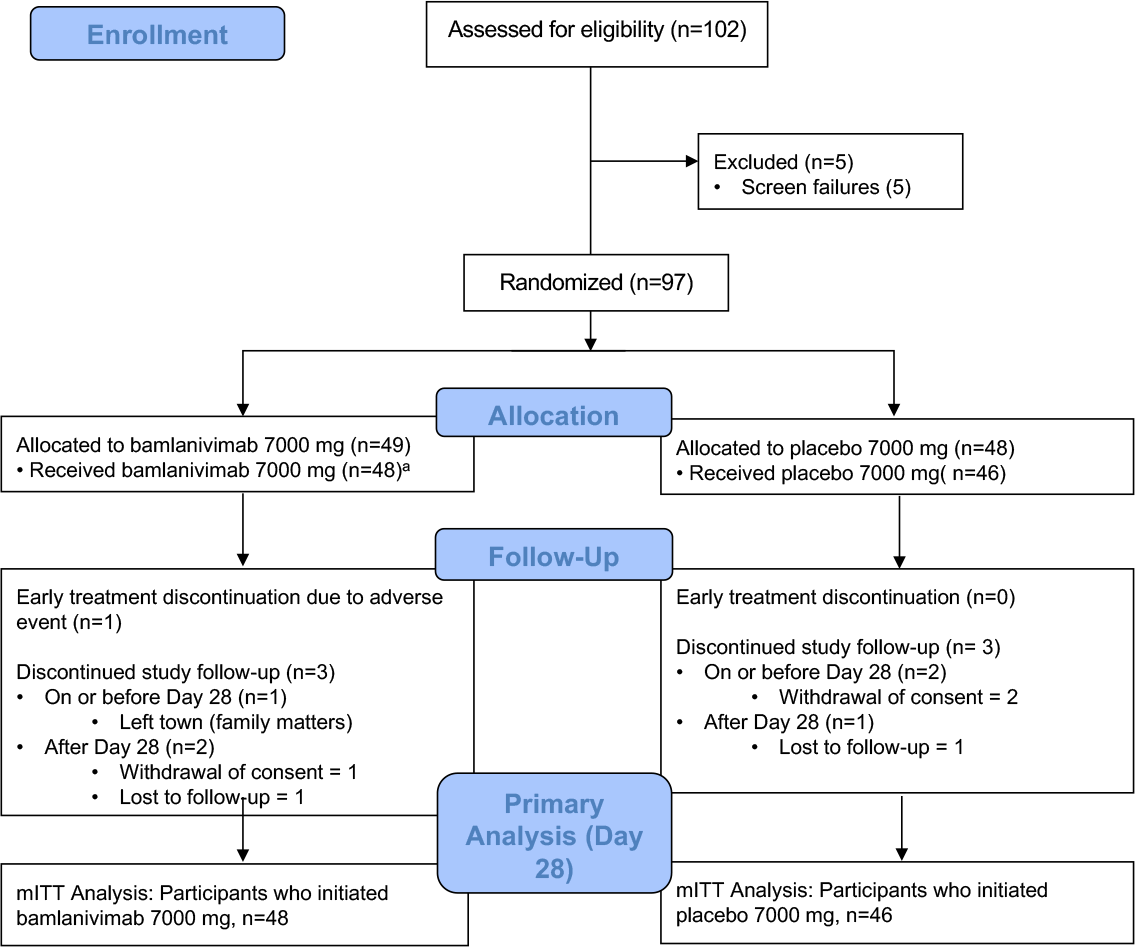
CONSORT Flow Diagram, Bamlanivimab 7000 Dose Group. ^a^Includes 1 participant assigned bamlanivimab 700 mg

**Figure 1B.**
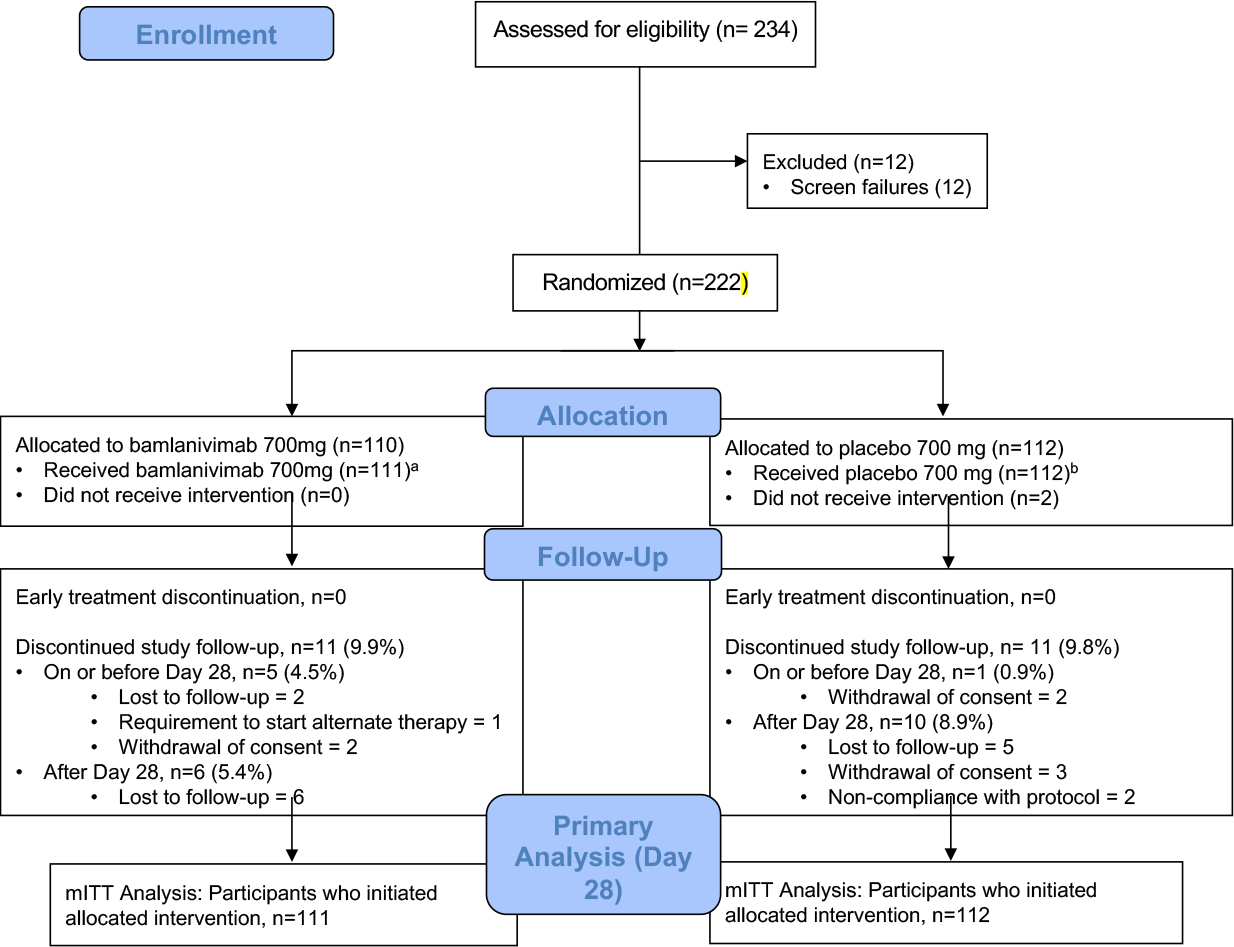
CONSORT Flow Diagram, Bamlanivimab 700 Dose Group. ^a^Includes 2 participants assigned bamlanivimab 7000 mg; ^b^Includes 2 participants assigned placebo 7000 mg

Participant characteristics were balanced across randomized arms in both the 7000 and 700 mg groups (**Table 1**). Across all 317 participants included in analyses, 116 (37%) reported ≤5 days of symptoms and 153 (48%) met the protocol definition of “higher” risk of progression to severe COVID-19. Hypertension, diabetes, obesity, and age were the most common high-risk criteria (**Supplementary Table 1**). The most frequently reported symptoms (reported by >40% of participants) within 48 hours of study entry included cough, headache, body pain or muscle pain/aches, fatigue, nasal obstruction or congestion, nasal discharge, and loss of taste or smell (**Supplementary Table 2**); most symptoms were reported as mild or moderate.

**Table 1.**
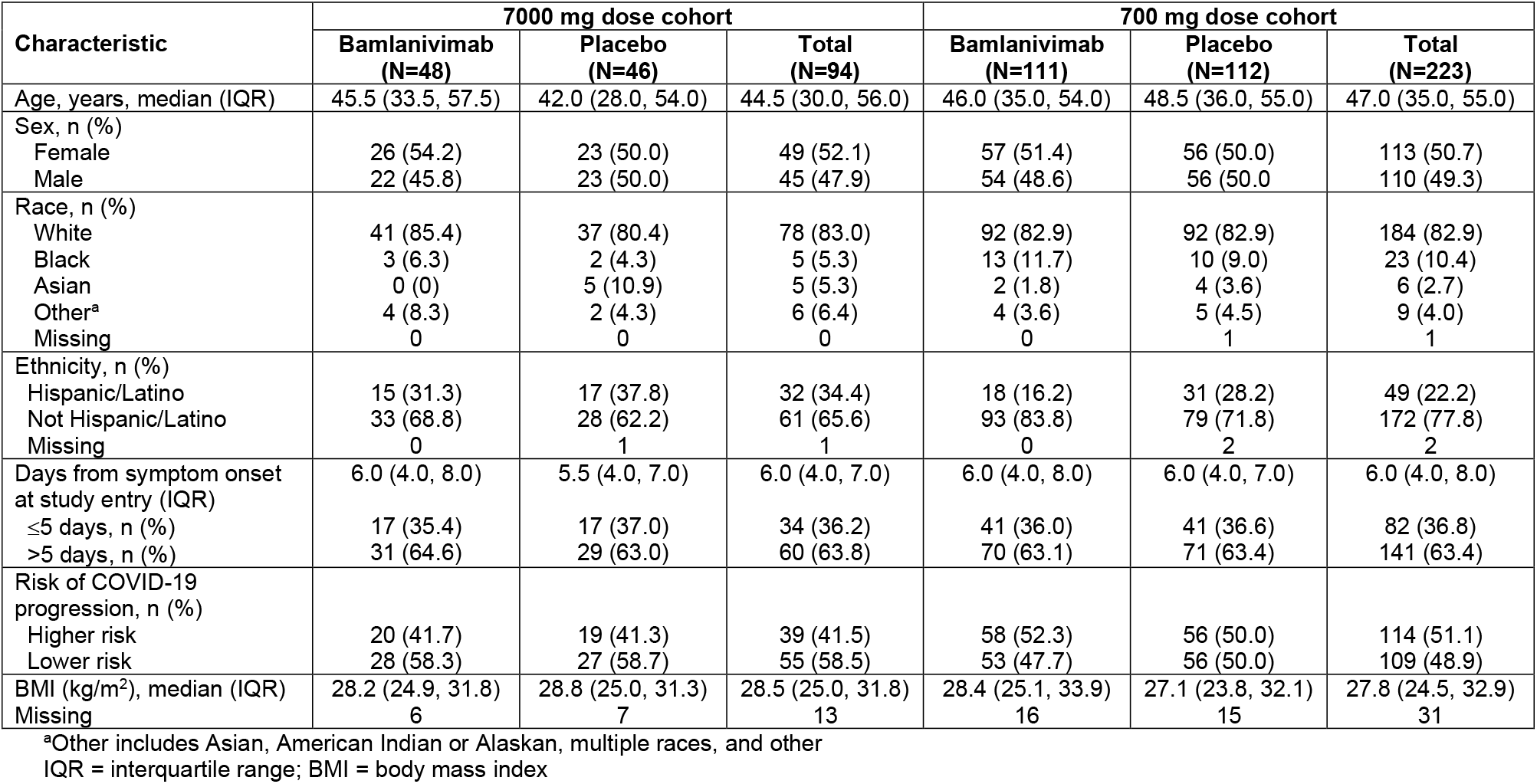
Baseline participant characteristics by dose group and treatment arm.

### Virological and Related Outcomes

Baseline NP and AN SARS-CoV-2 RNA levels (viral load) were highly correlated (ρ=0.85, p<0.001) (**Figure 2A**), with AN viral load lower than NP viral load (median [interquartile range, IQR) 4.5 [2.4, 6.3] and 5.7 [3.95, 6.8] log_10_ copies/mL for AN and NP swabs, respectively). Baseline NP and AN viral loads did not differ by risk category for COVID-19 progression (median [IQR] NP viral load 5.4 [3.5, 8.4] vs 5.5 [3.7, 6.6] log_10_ copies/mL, p=0.8) (**Figure 2B**), but viral loads were higher among participants entering the study with ≤5 days vs >5 days of symptoms (median NP viral load [IQR] 6.4 [5.2, 7.6] vs 4.8 [3.2, 6.1], p<0.0001) (**Figure 2C**) and among plasma SARS-CoV-2 viremic vs aviremic participants (median NP viral load [IQR] 6.4 [5.3, 7.6) vs 5.3 (3.3, 6.5), p<0.0001) (**Figure 2D**). Twenty percent of participants had detectable plasma SARS-CoV-2 RNA at baseline, without difference in proportion viremic by risk category for COVID-19 progression (**Figure 2E**). Total symptom score was higher among viremic vs aviremic participants at baseline (**Figure 2F**); symptom score did not correlate with NP or AN viral load (**Figure 2E**).

**Figure 2.**
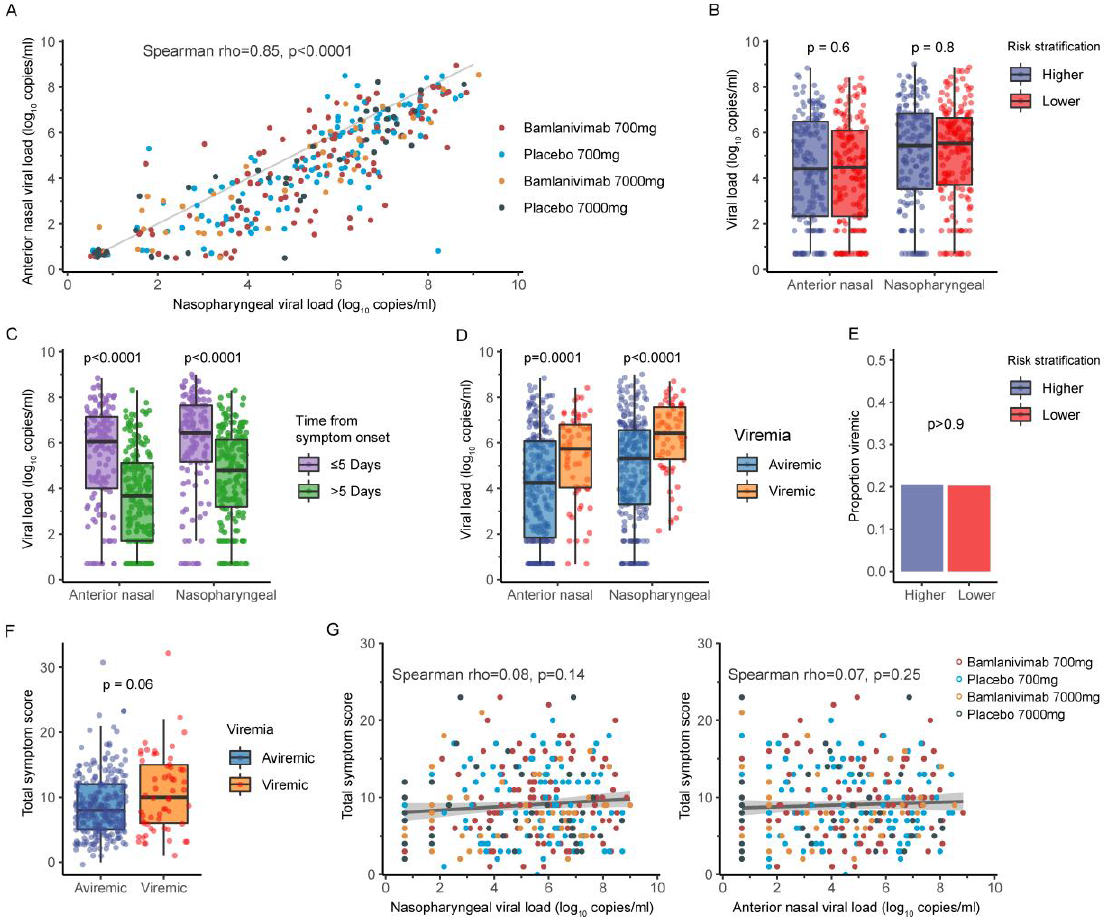
Associations between baseline virology and symptom scores and comparisons by subgroups, combining bamlanivimab 700 and 7000 mg dose cohorts. (A) Baseline nasopharyngeal (NP) and anterior nasal (AN) SARS-CoV-2 RNA levels (viral loads) were highly correlated. The diagonal line indicates line of equality. NP and AN viral loads did not differ by protocol-defined risk category (“higher” vs “lower”) for COVID-19 progression (B), but were significantly higher among participants entering the study with fewer days of symptoms (≤5 vs >5 days) (C) and among viremic vs aviremic participants (D). The proportion with SARS-CoV-2 viremia at study entry was the same for participants at higher vs lower risk for COVID-19 progression (E). Total symptom scores reported at study entry in the participant diary were higher among viremic participants (F). Symptoms scores did not correlate with NP or AN viral loads (G). In Figures 2B-2D and 2F, Tukey boxplots were used to demonstrate the distribution of viral loads or symptom score. Wilcoxon rank sum test was used to compare values from different groups.

Baseline NP SARS-CoV-2 RNA levels were similar at study entry between bamlanivimab vs placebo arms in each bamlanivimab dose cohort (**Supplementary Table 3**). The proportion of participants with undetectable NP SARS-CoV-2 RNA (primary virologic outcome) increased over time and did not differ between bamlanivimab or placebo arms for either dose cohort (**Figure 3 and Supplementary Table 3)**. At day 3, the median NP SARS-CoV-2 RNA level was significantly lower among bamlanivimab 700 mg recipients compared to placebo (2.9 vs 3.9 log_10_ copies/mL, p=0.002), and a similar trend was observed for bamlanivimab 7000 mg compared to placebo (2.2 vs 3.4 log10 copies/mL, p=0.07) **(Supplementary Table 3**). No differences in SARS-CoV-2 RNA levels between bamlanivimab and placebo groups were observed at any of the later visits (**Figure 3 and Supplementary Table 3**). Additionally, the AUC for SARS-CoV-2 RNA from day 0 through day 28 was smaller for both bamlanivimab 700 mg and 7000 mg compared to placebo, but neither difference met statistical significance (**Supplementary Table 3**).

**Figure 3.**
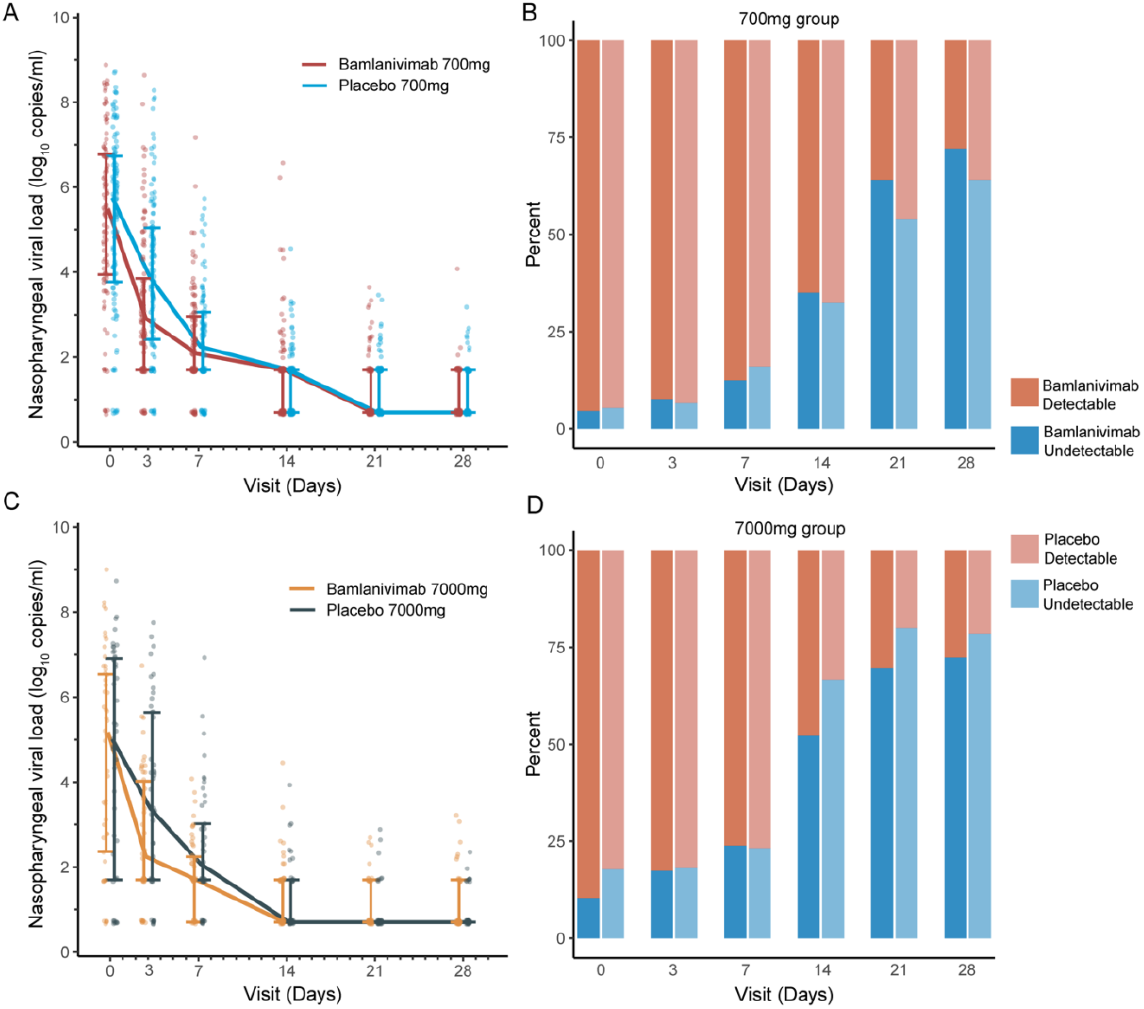
Nasopharyngeal (NP) SARS-CoV-2 RNA levels (viral loads) by dose cohort, treatment arm and visit. NP viral loads declined in all participants, with median NP viral loads lower at day 3 for bamlanivimab 700 mg (n=111) vs placebo (n=112) (2.9 vs 3.9 log_10_ copies/mL, p=0.002) (A), without a difference in proportion undetectable at any time points (B). Similar findings were seen, though differences in median viral load at day 3 were not statistically significant for the smaller 7000 mg dose cohort (n=48 bamlanivimab vs n=46 placebo, 2.2 vs 3.4 log10 copies/mL, p=0.07) (C and D). The lower limit of detection was 1.4 log_10_ copies/mL.

The viral load decay from NP and AN swab data was fitted for those participants for whom there was enough data (**Supplementary Table 4A)**. Decay rates were similar for each dose cohort (**Supplementary Table 4B**); thus, the 700 and 7000 mg dose cohorts were combined and fitted together, with separate analyses for NP and AN data. Population parameter estimates for the viral load decay in NP and AN swabs are provided in **Supplementary Table 4C**. The best model for AN data had a single exponential decay, and for the NP data, a biexponential decay. The first phase of viral decay was fast (AN: t_1/2_=7.8 and 6.5 hours and NP: t_1/2_= 10.3 and 7.2 hours for placebo and bamlanivimab-treated participants, respectively), while the second phase was slightly slower with t_1/2_=15.1 hours (in NP), with no difference in the second phase decay rate between placebo and bamlanivimab-treated participants. In both AN and NP models, the first phase of decay was significantly faster (p=0.0049 and p=0.0002, respectively) for bamlanivimab treatment compared to placebo.

### Symptoms and Other Clinical Outcomes

Overall, time to symptom improvement (primary symptom outcome) was long and did not differ significantly between bamlanivimab vs placebo arms for either dose cohort (median of 21 and 18.5 days for bamlanivimab 7000 mg vs placebo, p=0.97 and 24 vs 20.5 days for bamlanivimab 700 mg vs placebo, p=0.08) (**Table 2**). AUC of the total symptom score reported daily days 0-28 in the study diary also did not differ significantly between bamlanivimab vs placebo arms for either dose cohort (**Table 2**).

**Table 2.** Symptom outcomes by bamlanivimab dose cohort and treatment arm.

|  | 7000 mg dose cohort |  |  | 700 mg dose cohort |  |  |
| --- | --- | --- | --- | --- | --- | --- |
|  | Bamlanivimab<br>(N=48) | Placebo<br>(N=46) | p-value | Bamlanivimab<br>(N=111) | Placebo<br>(N=112) | p-value |
| Time to symptom improvement from study entry (primary symptom outcome), median (IQR), days | 21.0 (7.0, 28.0) | 18.5 (7.0, 28.0) | 0.97<sup>a</sup> | 24.0 (14.0, 28.0) | 20.5 (9.0, 28.0) | 0.08<sup>a</sup> |
| Proportion of participants with at least 1 symptom reported as more severe than at study entry in study diary, n (%) | 42 (87.5) | 40 (87.0) | 0.94<sup>b</sup> | 102 (91.9) | 105 (93.8) | 0.59<sup>b</sup> |
| Symptom severity ranking (AUC of total symptom score days 0-28), median (IQR) | 1.38 (0.93, 3.09) | 1.88 (1.09, 3.05) | 0.14<sup>a</sup> | 2.34 (1.30, 3.93) | 2.13 (1.06, 4.08) | 0.65<sup>a</sup> |

IQR = interquartile range, <sup>a</sup>Wilcoxon test, <sup>b</sup>chi-square test

CRP, ferritin, and fibrinogen levels declined more rapidly (greater fold change from baseline) in bamlanivimab 700 mg compared to placebo recipients at days 7 and 14 (as well as week 24 for CRP) **(Supplementary Figures 3 and 4)**. Greater fold-change reductions in prothrombin time (PT) were also observed at Days 14, 21, and Week 12 with bamlanivimab 700 mg. Similar trends were observed at some time points for bamlanivimab 7000 mg vs placebo (**Supplementary Figure 4**). No differences between bamlanivimab vs placebo arms were observed for fold change from baseline for LDH or activated PTT through day 28 (**Supplementary Figures 3 and 4**).

Through study day 28, there were 6 hospitalizations in the 7000 mg group, 2 (4.2%) on bamlanivimab and 4 (8.7%) on placebo, and 8 hospitalizations in the 700 mg group, 4 (3.6%) on bamlanivimab and 4 (3.6%) on placebo. No deaths were observed through week 24. Hospitalizations and deaths through week 24 are summarized in **Supplementary Table 5**.

### Safety

TEAEs through study day 28 are summarized in **Table 3** and TEAEs through week 24 in **Supplementary Table 6**. Grade 2 or higher and grade 3 or higher TEAEs were generally more frequently reported in bamlanivimab 700 and 7000 mg recipients than in placebo recipients, but the proportion with grade 3 or higher TEAEs (the primary safety outcome) did not differ significantly between bamlanivimab vs placebo arms for either dose and the vast majority of TEAEs were not felt to be related to study intervention. AESIs were infrequent and led to premature treatment discontinuation in only one participant assigned bamlanivimab 7000 mg, who did not complete the infusion due to a grade 3 IRR. SAEs through day 28 occurred in 2 (4.2%) and 4 (8.7%) of bamlanivimab 7000 mg and placebo recipients, respectively, and in 4 (3.6%) and 3 (2.7%) of bamlanivimab 700 mg and placebo recipients, respectively (**Table 3**). Detailed summaries of grade 2 and higher and grade 3 and higher TEAEs through day 28 by dose cohort and treatment arm are provided in **Supplementary Tables 7 and 8**.

**Table 3.** Adverse events (AEs) through day 28.

| Event | 7000 mg dose group |  |  | 700 mg dose cohort |  |  |
| --- | --- | --- | --- | --- | --- | --- |
|  | Bamlanivimab<br>(n=48) | Placebo<br>(n=46) | Risk Ratio<br>(bamlanivimab<br>7000 mg vs<br>placebo) (95%<br>CI), p-value<sup>a</sup> | Bamlanivimab<br>(n=111) | Placebo<br>(n=112) | Risk Ratio<br>(bamlanivimab<br>700 mg vs<br>placebo) (95%<br>CI), p-value<sup>a</sup> |
| Grade 3 or higher<br>TEAEs through day<br>28 (primary safety<br>outcome), number<br>of participants (%) | 6 (12.5) | 6 (13.0) | 0.96 (0.33,<br>2.76), p=0.94 | 10 (9.0) | 6 (5.4) | 1.68 (0.63,<br>4.47), p=0.30 |
| Grade 2 or higher<br>TEAEs through day<br>28, number of<br>participants (%) | 18 (37.5) | 14 (30.4) | 1.23 (0.697,<br>2.2), p=0.47 | 41 (36.9) | 25 (22.3) | 1.65 (1.08,<br>2.52), p=0.02 |
| AEs leading to<br>premature treatment<br>discontinuation,<br>number of<br>participants (%) | 1 (2.1) | 0 | -- | 0 | 0 | -- |
| AESIs through day<br>28, number of<br>participants (%) | 1 (2.1) | 2 (4.3) | -- | 1 (0.9) | 3 (2.7) | -- |
| Infusion-related<br>reaction | 1 (2.1) | 1 (2.2) | -- | 1 (0.9) | 1 (0.9) | -- |
| Hypersensitivity<br>reaction | 0 | 1 (2.2) | -- | 0 | 2 (1.8) | -- |
| Serious adverse<br>events (SAEs)<br>through day 28,<br>number of<br>participants (%) | 2 (4.2) | 4 (8.7) | -- | 4 (3.6) | 3 (2.7) | -- |

TEAE = treatment emergent adverse event; AESI = adverse event of special interest; <sup>a</sup>Wald test

### Pharmacokinetics

PK data were obtained on a total of 108 participants (71 who received 700 mg and 37 who received 7000 mg), as summarized in **Supplementary Table 9**. Mean Cmax values for the 700 mg and 7000 mg doses were 206 and 1867 µg/mL, respectively, and mean day 28 concentrations were 29 and 236 µg/mL, respectively. There was evidence for approximate dose proportionality, with geometric mean ratios for Cmax and AUC_0-∞_ of 8.3 and 8.5, respectively; the geometric mean ratio for total body clearance (CL) was 1.2. Interpatient variability was modest with coefficients of variation (CV) on CL of 40.5% at the 700 mg dose and 88.9% at the 7000 mg dose. Of bamlanivimab 700 mg recipients, 70/71 (98.6%) had day 28 concentrations above the estimated 90% inhibitory concentration (IC90) of bamlanivimab for SARS-CoV-neutralization of 4.2 μg/mL;^10^ one participant had bamlanivimab concentrations below the limit of quantitation at day 28.

## DISCUSSION

We present results of a phase 2 evaluation of the safety and efficacy of single dose bamlanivimab 700 mg and partially enrolled phase 2 evaluation of single dose bamlanivimab 7000 mg given by IV infusion for the treatment of non-hospitalized adults with COVID-19. For both bamlanivimab dose cohorts, in which participants received study intervention a median of 6 days from symptom onset, the intervention was safe, but no improvement was observed with bamlanivimab in the primary outcomes of proportion of participants with undetectable SARS-CoV-2 RNA from NP swabs or time to improvement of COVID-19-related symptoms. However, quantitative SARS-CoV-2 RNA levels from NP swabs were significantly lower among bamlanivimab-treated participants at a dose of 700 mg compared to placebo at day 3 of study, with a similar trend in the smaller 7000 mg dose cohort. Viral decay rates were modeled to be significantly faster for bamlanivimab compared to placebo. These observations are consistent with improvements in the semi-quantitative SARS-CoV-2 RNA measures by cycle threshold (Ct) observed with bamlanivimab in the BLAZE-1 trial. Of note, the SARS-CoV-2 RNA assay used in ACTIV-2/A5401 was fully quantitative and timing of treatment initiation after symptom onset was later than in BLAZE-1 (median of 6 vs 4 days of symptoms).^2^

Consistent with the reductions in NP SARS-CoV-2 RNA levels, we also found biological evidence of activity of bamlanivimab against COVID-19 progression, with greater reductions in inflammatory biomarker levels (CRP, ferritin, and fibrinogen) with bamlanivimab compared to placebo. While no impact of bamlanivimab therapy on symptom duration was found in our study, we note that symptom-based outcome measures for assessing treatment response have not yet been validated and persistence or brief recurrence of mild symptoms may have made our primary symptom outcome definition overly sensitive to symptoms that may not have been clinically meaningful. Across outpatient COVID-19 therapeutic studies, definitions of symptom improvement or resolution, symptom diaries, severity scoring scales. analytical approaches, and included symptoms have differed. The BLAZE-1 study first examined change from baseline in symptom scores and found modest early reductions in symptom scores with bamlanivimab compared to placebo.^2^ Subsequent analyses of the combination mAbs bamlanivimab and etesevimab examined different definitions of symptom resolution or improvement, and found a 1 day reduction (8 vs 9 days) in median time to sustained resolution of symptoms, where sustained resolution of symptoms was defined by 2 consecutive days of absent symptoms (6 targeted), allowing for ongoing mild cough and fatigue.^11^

The combination mAbs casirivimab and imdevimab reduced symptom duration by 4 days (from 14 to 10) utilizing a different symptom resolution definition of time to first day the participant scored 19 symptoms absent, allowing ongoing “mild/moderate” cough, fatigue, and headache, and restricting to participants with a minimum symptom score >3.^12^ One obvious impact of the different definitions of symptom resolution is on the duration of symptoms from study entry – shorter durations were reported in the BLAZE-1 and REGEN-COV studies than in our study (median durations of >20 days). Based on clinical observations that COVID-19 symptoms may wax and wane day-to-day, we sought to minimize the possibility of recurrent or relapsing symptoms in defining our population with improved symptoms, resulting in long durations of symptoms. An additional challenge is distinguishing between symptoms that are specific to COVID-19 or due to comorbidities, given many viral illness symptoms are non-specific. The potential for mAbs to modify symptom duration and accelerate symptom resolution may also depend on timing of mAb therapy during the COVID-19 disease course – where later administration (as late as 10 days after symptom onset, as in our study and consistent with current mAb EUA guidance) may have less impact, although this is unknown. Validation of COVID-19 symptom diary content and outcome measures are ongoing, by our group and others.

Additionally, the impact of early antiviral therapy and mAbs on long-term post-acute sequelae of SARS-CoV-2 infection (PASC) is unknown. Defining and measuring PASC presents similar, if not greater challenges to those of defining acute COVID-19 symptom resolution/improvement. The reductions in inflammatory biomarkers with bamlanivimab, including as late as 24 weeks after treatment, suggest some promise for early antiviral therapy to mitigate or prevent the development of PASC, or some PASC presentations. Studies to quantify the potential benefit of early antiviral therapy on late COVID-19 sequelae are also ongoing, by our group and others.

The ACTIV-2/A5401 outcome measures were designed early in the COVID-19 pandemic, and our understanding of the SARS-CoV-2 viral kinetics on NP and AN sampling continues to evolve. At the time the study was designed, the dynamics of RNA shedding from the nasopharynx and anterior nares in people with mild-moderate symptoms and correlates of NP and AN viral levels were not well described. Our study clearly demonstrates that while viral shedding in these compartments was not greater in participants at higher risk of disease progression or correlated with self-reported symptom scores, it was associated with shorter duration of symptoms and NP (and AN, data not shown) RNA shedding declines rapidly in most individuals, even in the absence of treatment. NP RNA shedding as a measure of antiviral activity is likely to be most informative early after symptom onset and with very early sampling following treatment.

Consistent with this, greater reductions in NP SARS-CoV-2 RNA compared to placebo have been observed for multiple anti-SARS-CoV-2 neutralizing mAbs in parallel with reductions in COVID-19-related hospitalizations and deaths, in studies where the mAbs were administered early after symptom onset (median of 3-4 days), and earlier than in our study.^2-4,11^ Whether antiviral/mAb therapy has a clinical benefit after a longer duration of symptoms is a separate question from detecting antiviral activity (i.e. the value of change in NP RNA levels as a surrogate for clinical outcomes is likely limited to a short window during early infection, when RNA levels are high, and may not be similarly predictive or correlated later in the disease course, nor reflective of antiviral activity). Indeed, our group has demonstrated the clinical benefit of anti-SARS-CoV-2 mAbs (the combination of BRII-196 and BRII-198 administered IV) given as late as 10 days after symptom onset, with similar reductions in hospitalizations and deaths when given >5 days vs within 5 days from symptom onset;^13^ analyses of NP RNA shedding and hospitalization/death rates by symptom duration at time of treatment in this phase 2/3 trial within the ACTIV-2/A5401 platform will be informative.

Our study also demonstrates that NP and AN RNA shedding is greater among non-hospitalized persons with SARS-CoV-2 viremia compared to those without detectable viremia, and symptom scores also tend to be higher among viremic compared to aviremic participants. Given the small number of hospitalizations, we could not determine if nasal shedding, viremia, or changes with treatment were associated with this outcome or COVID-19 severity, as has been observed in hospitalized persons.^14,15^

In this study, we also expand on the reported PK of bamlanivimab and found the characteristics in 108 non-hospitalized persons were comparable with those reported in the first-in-human study in hospitalized patients for both the 700 and 7000 mg doses.^16^ Absolute dose proportional PK were not observed, explained by the faster total body clearance with 7000 mg than 700 mg. The 700 mg dose (the dose that was granted an EUA) achieved sustained serum concentrations above the predicted IC_90_ for SARS-CoV-2 neutralization in nearly all participants with available PK data, supporting the selection of this dose for clinical use. However, bamlanivimab concentrations in target tissues such as the respiratory tract have not been measured in humans and whether this same pharmacodynamic relationship exists in tissues is not known.

These phase 2 trial results affirm the overall safety and antiviral activity of bamlanivimab reported previously and suggest a benefit on systemic inflammation. While the safety and efficacy of anti-SARS-CoV-2 mAbs for the prevention of hospitalizations and deaths in persons at high risk of progressive disease have been established, further evaluation is needed to define the benefit of early treatment with mAbs on symptom outcomes in persons at lower risk for severe COVID-19 and on longer-term outcomes, such as PASC.

## Data Availability

The authors confirm that all data underlying the findings are fully available. Due to ethical restrictions, study data are available upon request from with the written agreement of the AIDS Clinical Trials Group and the manufacturer of the investigational product.

## Acknowledgements

We thank the AIDS Clinical Trials Group, including Lara Hosey and Jhoanna Roa; the National Institute of Allergy and Infectious Diseases (NIAID) / Division of AIDS (DAIDS), including Peter Kim; the U.S. Government Response to COVID-19, including Bill Erhardt; the Foundation for the National Institutes of Health and the Accelerating COVID-19 Therapeutic Interventions and Vaccines (ACTIV) partnership, including Stacey Adams; and PPD.

We also thank the members of the ACTIV-2/A5401 data and safety monitoring board — Graeme A. Meintjes, PhD, MBChB (Chair), Barbara E. Murray, MD, Stuart Campbell Ray, MD, Valeria Cavalcanti Rolla, MD, PhD, Haroon Saloojee, MB.BCh, FCPaed, MSc, Anastasios A. Tsiatis, PhD, Paul A. Volberding, MD, Jonathan Kimmelman, PhD, David Glidden, PhD, and Sally Hunsberger, PhD (Executive Secretary).

## Funding

This work was supported by the National Institute of Allergy and Infectious Diseases of the National Institutes of Health under Award Number 3UM1AI068636-14S2, UM1 AI068634, UM1 AI068636 and UM1 AI106701. The content is solely the responsibility of the authors and does not necessarily represent the official views of the National Institutes of Health. Portions of this work were performed at Los Alamos National Laboratory under the auspices of the US Dept. of Energy under contract 89233218CNA000001 and supported by NIH grants R01-AI028433, R01-OD011095 (ASP), and Los Alamos National Laboratory LDRD 20200743ER, 20200695ER, and 20210730ER. Study medication was donated by Eli Lilly and Company.

## Conflicts of Interest

KWC has received research funding to the institution from Merck Sharp & Dohme. ESD receives consulting fees from Gilead Sciences and Merck and research support through the institution from Gilead Sciences and ViiV. DAW has received funding to the institution to support research and honoraria for advisory boards and consulting from Gilead Sciences. JZL has consulted for Abbvie. ASP has consulted for Amphylx Pharmaceticals. C. Malvestutto has received research funding to the institution from Eli Lilly. PK, KP, and AN are employees and shareholders of Eli Lilly. WF has received research funding from Ridgeback Biopharmaceuticals, served on adjudication committees for Janssen, Syneos, and consulted for Roche and Merck. JJE is an ad hoc consultant to GSK/VIR, data monitoring committee (DMC) chair for Adagio Phase III studies. DMS has consulted for the following companies Fluxergy, Kiadis, Linear Therapies, Matrix BioMed, Arena Pharmaceuticals, VxBiosciences, Model Medicines, Bayer Pharmaceuticals, Signant Health and Brio Clinical.

## Notes

### Clinical Trial

NCT04518410

### Author Declarations

Advarra IRB gave ethical approval for this work (Pro00045266).

